## Appendix for "Programmatic considerations for chikungunya vaccine introduction in countries at risk of chikungunya outbreaks: stakeholder analysis"

**Appendix Table 1.** Interview questionnaire used in the semi-structured interviews. These questions were used to guide the interviews, but participants were invited to speak freely and structure the interview how they saw best.

| **#** | **Question** |
| --- | --- |
| Q1 | Please introduce yourself, explain your role within your organisation and summarise your work related to chikungunya |
| Q2 | Based on your experience, what is your perception of (your region's) current risk of a chikungunya outbreak? |
| Q3 | Do you think a chikungunya vaccine (in your region) would be feasible? |
| Q4 | What are the potential barriers in chikungunya vaccine uptake? |
| Q4b | If not covered in Q4, what political, social, financial, logistical barriers would affect the roll-out of chikungunya vaccine in (your area)? What other factors would affect the feasibility of vaccination? |
| Q5 | Is there any other information you think would be useful for us to know? Is there anything else unique to your experience in your region that you would like to share? |
